# Targeted gene expression profiling for accurate endometrial receptivity testing

**DOI:** 10.1101/2022.06.13.22276318

**Authors:** Alvin Meltsov, Merli Saare, Hindrek Teder, Priit Paluoja, Riikka Arffman, Terhi Piltonen, Piotr Laudanski, Mirosław Wielgoś, Luca Gianaroli, Mariann Koel, Maire Peters, Andres Salumets, Priit Palta, Kaarel Krjutškov

**Affiliations:** Competence Centre on Health Technologies; Tartu, Estonia; Department of Genetics and Cell Biology, GROW School for Oncology and Developmental Biology, Maastricht University, Maastricht, The Netherlands; Department of Obstetrics and Gynecology, Institute of Clinical Medicine, University of Tartu; Tartu, Estonia; Institute of Biomedicine and Translational Medicine, University of Tartu; Tartu, Estonia; Department of Obstetrics and Gynecology, PEDEGO Research Unit, Medical Research Center, Oulu University Hospital, University of Oulu; Oulu, Finland; Department of Obstetrics and Gynecology, Medical University of Warsaw; Warsaw Poland; Oviklinika Infertility Center; Warsaw, Poland; SISMeR, Reproductive Medicine Unit; Bologna, Italy; Institute of Genomics, University of Tartu; Tartu, Estonia; Division of Obstetrics and Gynecology, Department of Clinical Science, Intervention and Technology (CLINTEC), Karolinska Institutet and Karolinska University Hospital; Stockholm, Sweden; Activate Health, Tallinn, Estonia; Institute for Molecular Medicine Finland (FIMM), University of Helsinki; Helsinki, Finland

**Keywords:** endometrium, endometrial receptivity, beREADY test, TAC-seq, window of implantation, personalised embryo transfer

## Abstract

**STUDY QUESTION:** How accurately can a targeted gene expression sequencing assay estimate endometrial receptivity corresponding to the window of implantation?

**DESIGN:** Endometrial biopsies (n=175) from healthy fertile volunteers (n=66), polycystic ovarian syndrome (PCOS) patients (n=39), and recurrent implantation failure (RIF) patients (n=44) were collected and sequenced with TAC-seq (Targeted Allele Counting by sequencing) method targeting 68 biomarker genes for endometrial receptivity. The expressional profiles were clustered, and differential expression analysis was performed on the model development group, using 63 endometrial biopsies spanning over proliferative (PE, n=18), early-secretory (ESE, n=18), mid-secretory (MSE, n=17) and late-secretory (LSE, n=10) endometrial phases. A quantitative predictor model was trained on the development group and validated on sequenced samples from healthy women, consisting of 52 paired samples taken from ESE and MSE phases and five LSE phase samples from 31 individuals. Finally, the developed test was applied to 44 MSE phase samples gathered from a study group of patients diagnosed with RIF.

**RESULTS:** The developed assay successfully captures the unique receptivity profile of the endometrium using 68 biomarker genes. When compared to healthy women of the same cycle phase, we did not detect any significant gene expression bias caused by PCOS in PE, ESE, MSE, and LSE samples. In validation samples (n=57), we detected displaced WOI in 1.8% of the samples from fertile women. In the RIF study group, we detected a significantly higher proportion of the samples with shifted WOI than in the validation set of samples from fertile women, 15.9% and 1.8%, respectively.

**CONCLUSIONS:** The developed beREADY screening test enables highly sensitive and dynamic detection of selected transcriptome biomarkers, providing a quantitative and accurate prediction of endometrial receptivity status for personalised embryo transfer.

## Introduction

Infertility affects millions of people of reproductive age worldwide, and increasingly more couples undergo in vitro fertilisation (IVF) to achieve pregnancy [1]. Though IVF success rates have improved significantly, many patients still fail to conceive and experience recurrent implantation failure (RIF). The reasons for embryo implantation failure are highly heterogeneous, attributable to the quality of the embryo, the endometrium, and the interaction between the two [2]. As these circumstances vary from patient to patient, applying a personalised approach for assessing both embryo quality and endometrial receptivity could increase the chance of implantation during IVF.

The period when the endometrium becomes receptive for embryo implantation is called the window of implantation (WOI). There is no consensus about the exact timing and length of the WOI period. Generally, the endometrium is considered to become receptive seven days after the luteinising hormone (LH) peak during the natural cycle (NC) and lasts for two days. However, several studies suggest that the length of the WOI may vary from two to up to six days [3–6]. In addition, based on transcriptional studies of the endometrial biopsies, the WOI can be shifted or displaced in time [7, 8]. It has been estimated that displaced WOI occurs in around 10% of women undergoing IVF and at least 25% of women with RIF [6], [9–11]. In this light, the determination of the personalised WOI should be a standard procedure for women undergoing IVF to avoid implantation failure after embryo transfer due to the asynchrony between the endometrial and embryonal development. Therefore, this procedure may be particularly beneficial for RIF patients.

Polycystic ovary syndrome (PCOS) is an infertility-associated disorder common among women of reproductive age. It is still unclear whether endometrial receptivity is affected in PCOS patients. Dysregulation of several endometrial receptivity-associated genes in PCOS patients has been described (reviewed [12]). Besides the altered gene expression profile of endometrial tissue, PCOS patients are often affected by obesity, hyperinsulinemia, and increased general inflammation, likely contributing to infertility [13]. Nevertheless, to the best of our knowledge, no systematic studies have been published describing the gene expression signature of essential endometrial receptivity genes in PCOS throughout the entire menstrual cycle.

Considerable effort has been made to describe molecular changes in the menstrual cycle during WOI [14, 15]. The hormone-induced regulatory cascade leads to endometrial maturation and major changes in gene expression culminating with WOI. The transcriptomic landscape of endometrial maturation is well characterised through whole transcriptome studies where different gene expression profiles have been detected between proliferative (PE), early- (ESE), mid- (MSE), and late-secretory (LSE) endometrium [14, 16, 17]. Based on the studies, biomarkers for WOI determination and personalised embryo transfer (pET) have been provided. Diagnostic tests based on gene expression profiling of varying sets of endometrial receptivity biomarkers have been developed and integrated into infertility treatment [18-20]. Endometrial receptivity tests currently in clinical use are ERA® test (Igenomix) [18], ER Map® test (IGLS) [20], WIN-Test (INSERM) [21], and rsERT test (Yikon Genomics Company) [11]. Compared to the targeted sequencing approach used in the present study, these tests are limited either by the accuracy and dynamic range in detecting transcript abundances or by the scalability of the technique applied.

This study introduces the beREADY test for reliable WOI detection for pET. beREADY test was developed on Illumina sequencing-based TAC-seq technology (Targeted Allele Counting by sequencing), enabling biomolecule analysis down to a single-molecule level [22]. The 72 genes analysed with this test contain a core set of 57 endometrial receptivity-associated biomarkers [23], 11 additional genes, and four housekeeper genes.

## Materials and Methods

The study was approved by the Research Ethics Committee of the University of Tartu, Estonia (333/T-6) and by the Research Ethics Committee of the Hospital District of Northern Ostrobothnia, Finland (No 65/2017). Written informed consent was obtained from all participants prior to enrolling in the study.

### Patient selection and sample collection

Three different study groups were used for the beREADY test model training and testing. The general characteristics of these groups are presented in **Table 1**. The menstrual cycle phase was confirmed by menstrual cycle history and LH peak measurement using Clearblue® digital ovulation test or BabyTime hLH urine cassette (Pharmanova) and histological evaluation of biopsies according to Noyes’ criteria. None of the women had been using any hormonal medications for at least three months before the biopsy. All endometrial tissue biopsies were collected using a Pipelle® flexible suction catheter (Laboratoire CCD, France).

**Table 1.** General characteristics of study participants

|  | Model training and development group (MD) |  | Model validation group (MV) | RIF study group (RIF) |
| --- | --- | --- | --- | --- |
| Group description | Healthy (n=35) | PCOS (n=39) | Healthy (n=31)* | RIF (n=44) |
| Age (years, mean ± SD) | 34.1±3.7 | 34.5±3.6 | 30.1±3.1 | 35.9±3.9 |
| BMI (kg/m<sup>2</sup>, mean ± SD) | 23.1±2.5 | 24.7±3.2 | 23.1±4.3 | 23.6±3.7 |
| Proliferative (n) | 10 | 10 | - | - |
| Early-secretory (n) | 10 | 10 | 26 | - |
| Mid-secretory (n) | 10 | 10 | 26 | 44 |
| Late-secretory (n) | 5 | 9 | 5 | - |

BMI- body mass index, RIF- recurrent implantation failure, SD- standard deviation.
\* Paired samples obtained from the same NC in mid-secretory phase and early-secretory phase groups

#### Model training and development group

Endometrial samples in the model training and development group (MD) were collected from different menstrual cycle phases of the natural cycle (NC) from healthy fertile volunteers (n=35) and women with PCOS (n=39) at the Oulu University Hospital (Finland), **Table 1**. The BMI matched healthy controls for the PCOS group and had a regular menstrual cycle without any signs of PCOS or endometriosis. The women with PCOS were identified from the hospital register based on their prior diagnosis of PCOS. Their PCOS phenotype was confirmed by interview, clinical examination, and vaginal ultrasonographic examination of ovaries (Voluson 730 Expert). All women with PCOS diagnosis had polycystic ovarian morphology, current or previous oligomenorrhea, and two had signs of clinical hyperandrogenism. In healthy and PCOS groups, proliferative (PE), early-secretory (ESE), mid-secretory (MSE), and late-secretory (LSE) endometrial samples were collected.

#### Model validation group

The model validation group (MV) consisted of 26 pairs of endometrial tissue samples from healthy volunteers at ESE and MSE phases. In addition, we included endometrial tissue samples collected from five healthy volunteers at LSE phase. Therefore, we sequenced 57 endometrial tissue samples from 31 volunteers (**Table 1**). The paired NC ESE and MSE samples were collected at the Nova Vita Clinic (Tallinn, Estonia), and the LSE samples were collected from Oulu University Hospital (Finland). All women had normal serum levels of progesterone, prolactin, and testosterone, negative screening results for sexually transmitted diseases, no uterine pathologies, no signs of endometriosis or PCOS, and at least one live-born child. Tissue histology evaluated one MSE phase tissue sample belonging to PE phase, and one ESE sample had ambiguous histological morphology with minor similarities to the MSE samples. With these exceptions, the results of the histological evaluation were concordant with the time of the biopsy.

#### RIF study group

RIF group samples were obtained from women undergoing IVF at the Nova Vita Clinic in NC (n=44, **Table 1**). The endometrial biopsies were collected for research purposes, and no frozen embryos were replaced in the same cycle. All RIF patients had undergone, on average, 3.8 previous unsuccessful IVF cycles with good quality embryos, and the causes for infertility treatment were tubal (n=15), male (n=11), unknown (n=8), endometriosis (n=2), and other factors (n=8).

### RNA extraction from endometrial tissue

According to the manufacturer’s protocol, endometrial tissue total RNA was extracted using miRNeasy or RNeasy Mini kit (Qiagen). DNase I treatment was performed on column using RNase-Free DNase Set (Qiagen). Purified RNA integrity number (RIN) and quantity were assigned with Bioanalyzer 2100 RNA Nano 6000 kit (Agilent Technologies) and Qubit RNA IQ Assay (Thermo Fisher Scientific). Samples with RIN ≥ 7 were considered eligible for further analysis.

### Biomarker selection and assay design

In total, 57 previously published biomarkers were used as endometrial receptivity biomarker genes [23] together with four housekeeping genes *(SDHA, CYC1, TBP*, and *HMBS*) [22] and 11 additional genes (*CAMK2D, CAAP1, FOXN2, GGNBP2, ICA1L, LEFTY1, OGT, PPIP5K2, RIC3, TPM2*, and *YARS2*).

Highly sensitive and quantitative TAC-seq technology [22] was applied for endometrial receptivity biomarker profiling (**Fig. 1**). Briefly, the TAC-seq assay was modified to analyse mRNA biomarkers based on their oligo-T primed cDNA synthesis. The robustness of the screening test was increased by designing TAC-seq specific DNA oligonucleotides as probes to be located close to the biomarker’s mRNA 3’-end, enabling higher tolerance for RNA degradation. Each biomarker molecule was detected by stringent hybridisation of two specific DNA probes close to each other on the cDNA molecule. Once the DNA oligonucleotides are hybridised, the strands are joined enzymatically. The formed complex has all required components for further quantitative analysis, including unique molecular identifier (UMI) motifs (2×4bp UMI barcode per complex). The application of the UMI method allows for the identification and removal of PCR duplicates *in silico*, enabling the quantification of transcripts of interest at a single-molecule level.

**Figure 1.**
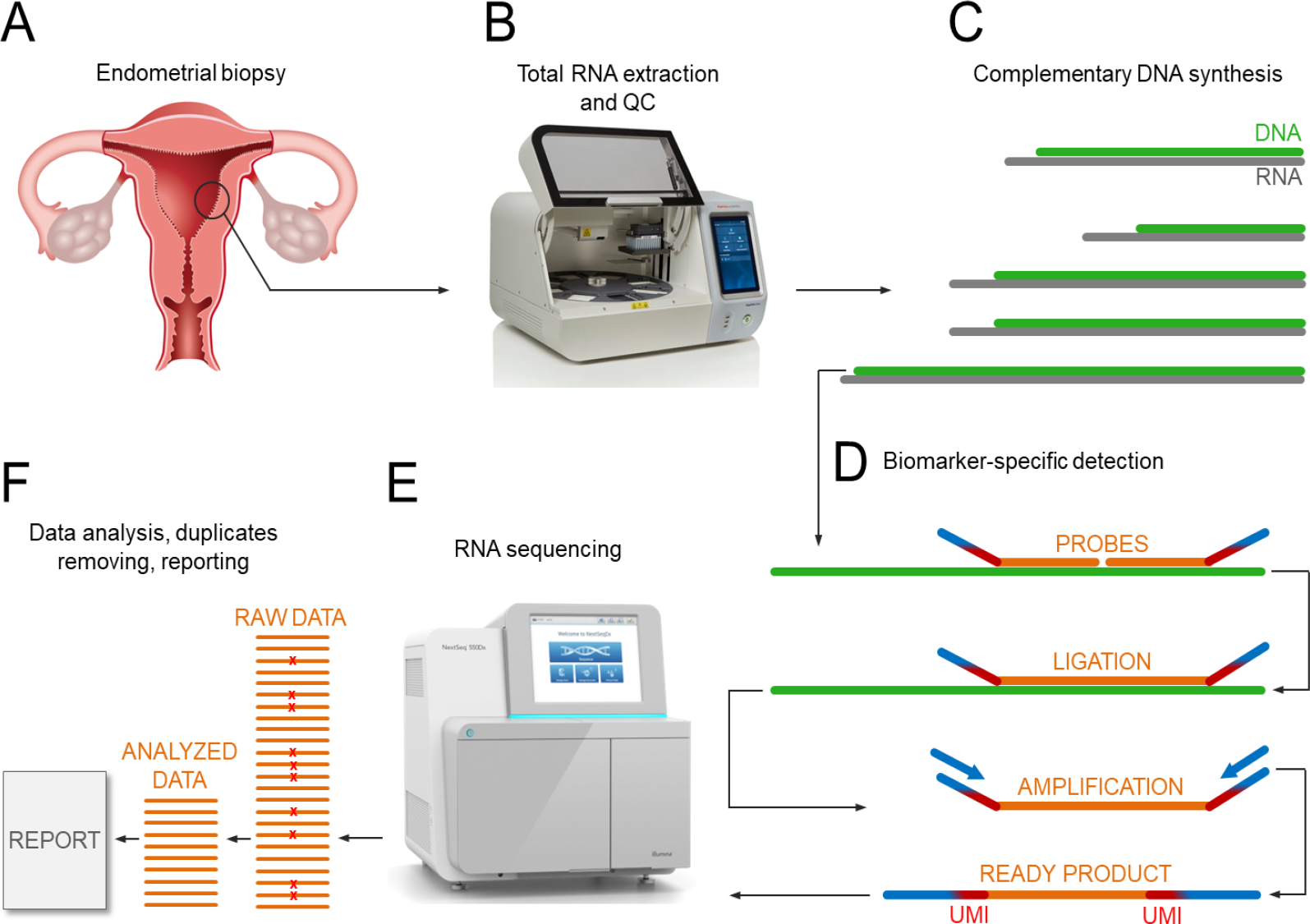
Illustrative schematic overview of beREADY endometrial receptivity biomarker analysis by TAC-seq technology. (A) Endometrial biopsy is the source of total RNA that contains a set of analysed biomarker genes. (B) The biopsy’s visual quality control, total RNA extraction, and quality control (QC). (C) The set of selected biomarkers’ RNA molecules in grey for the template of complementary DNA in green after oligo-T priming. (D) Biomarker-specific detection through DNA-DNA hybridisation with orange probes. The ligation joints the probe DNA molecules only in a perfect match. The blue arrows represent PCR primers introducing patient-specific barcodes that copy red UMI parts and orange biomarker regions for RNA sequencing. (E) Illumina RNA sequencing technology generates millions of reads per analysed sample. (F) The orange RNA sequencing reads represent raw data that includes technical PCR duplicates marked with red crosses. Further bioinformatic analysis recognises and removes the duplicates based on UMI-s and profiles the sample of interest naturally. The final step is a report.

### Library preparation and sequencing

The detailed protocol of TAC-seq was published previously [22], but critical updates were applied to develop the endometrial receptivity testing assay. For Illumina-compatible library preparation, 4 µl total-RNA (50 ng/µl) was first denatured 2 min at 80°C and then mixed with 1 µl FIREScript® RT cDNA synthesis MIX (Solis BioDyne, Estonia). One microliter of TAC-seq probe mixture was added to previously prepared cDNA. After an hour of stringent hybridisation at 60°C, the thermostable ligase was mixed according to the protocol. Further, PCR was performed in 40 µl volume containing 1× proofreading HOT FIREPol Blend Master Mix (Solis BioDyne) and 250 nM primers. PCR products were pooled, purified with DNA Clean & Concentrator-5 column kit (Zymo Research), and eluted with 50 µl of elution buffer (EB). Mag-Bind Total Pure NGS beads (Omega Bio-Tek) were added to 50 µl of the purified PCR product, incubated for 5 min at room temperature, and captured by a magnet for 3 min. After incubating, the supernatant was discarded, and beads were eluted in 25 µl of EB. The 180 bp uniform libraries were visualised and quantified on a TapeStation High Sensitivity D1000 ScreenTape (Agilent Technologies). High-coverage TAC-seq libraries were sequenced with Illumina NextSeq 550 high output 75 cycles kit.

### Sequencing data processing

TAC-seq sequencing data processing is described in detail [22], and open-source software for TAC-seq data processing is available at https://github.com/cchtEE/TAC-seq-data-analysis. Firstly, the software pipeline was used to match sequencing reads to the target sequences in the beREADY assay. Up to five mismatches per target sequence were allowed when matching barcodes. Next, UMI-thresholding was applied by merging target sequence counts with the same UMI sequence. For the beREADY sequencing assay, a UMI threshold of at least one unique UMI per target sequence was used. Finally, after obtaining the molecule count estimates, the counts were normalised considering the geometric mean of the molecule counts of the housekeeping genes. The resulting gene expression levels for each sample were used for further downstream analysis.

### Quality control in endometrial samples

The MD group included endometrial samples collected in different menstrual cycle phases from healthy fertile volunteers (n=35) and women with PCOS (n=39) at the Oulu University Hospital (Finland) (**Table 1**). For the PCOS analyses and beREADY model development, 11 endometrial samples were excluded due to insufficient sample quality after sequencing or unclear tissue histology. As a result, endometrial samples were used from 63 women (18 samples from PE, 18 from ESE, 17 from MSE, and 10 from the LSE phase).

### Statistical analysis

Statistical analysis, model development and visualisation were done in R language (v4.1.2) [24]. Before statistical testing, shifted logarithm transform was applied to the quantified and normalised read counts. The significance of the difference in proportions of displaced WOI in MV and RIF groups was tested with a lower-tailed Fisher’s exact test. In the case of comparative analysis between two groups, an independent t-test with Bonferroni correction was applied to find significant (p < 0.05) differentially expressed genes. A two-sided analysis of variance (ANOVA) was performed when investigating the interaction of two independent variables, and the interaction score was evaluated with the F-test.

For assessing the accuracy of the computational model, 5-fold cross-validation was applied. In every iteration, samples were randomly divided into five subgroups, out of which four were used to train the model and one to test. At least one random sample from each group was assigned to the testing group during the stratification. The test samples were classified with the most probable receptivity class by comparing every class’s relative receptivity probability outputs. This procedure was repeated 100 times, and the average accuracy over all the classes was reported.

### beREADY computational model development

The model for detecting endometrial receptivity was based on relative cluster distances to the MD phase groups after dimensionality reduction with principal component analysis (PCA). Briefly, the MD samples were normalised with the geometric mean of housekeeper gene expression levels and scaled. Next, Horn’s parallel analysis (100 iterations; 0.05 quantile) determined the number of principal components to keep [25]. The development set eigenvectors and scaling parameters from the PCA were used to project MV or RIF samples to the previously selected principal components. After projection, the centroids for the MD receptivity phases were calculated. Subsequently, squared Mahalanobis distances of the projected samples to the reference set group centroids were calculated. Samples with p-values corresponding to the lower tail of the χ2 distribution smaller than 0.025 were considered outliers [26]. This procedure was repeated for the closest pair of groups. The relative probability of the test sample belonging to either of the closest groups was reported. Only distances to the closest temporally adjacent receptivity stages were compared on the second iteration, constructing a hierarchical sequence of exclusive predictive classes. The relative receptivity class (‘pre-receptive’, ‘receptive’ and ‘post-receptive’) probabilities for each studied sample were reported.

## Results

### Study design

First, gene expression signatures of 68 endometrial receptivity genes were analysed in the MD group samples (n=63), consisting of both healthy volunteers and PCOS patients. Based on collected data, a continuous and quantitative three-stage (from pre-receptive to receptive to post-receptive) computational classification model was developed. Next, the model classification accuracy was validated on MV group samples (n=57, including ESE, MSE, and LSE samples). Finally, the RIF group samples (n=44, women with RIF in NC at MSE) were examined with the validated computational classification model. The outline of this study is presented in **Supplementary Figure 1**.

### Gene expression profiling of PCOS samples

This analysis of endometrial receptivity biomarkers revealed no difference between healthy women and PCOS patients (**Fig. 2**). Comparative t-testing and ANOVA of individual genes did not reveal any significantly differentially expressed genes (FDR≥0.05) between the groups of PCOS patients and healthy women (**Suppl. Table 1**). These results were observed in all four menstrual cycle phases (**Fig. 2**) and confirmed by principal component analysis (**Suppl. Fig. 2**). Based on these results, we concluded that PCOS status does not affect the expression profiles of biomarkers included in the developed assay.

**Figure 2.**
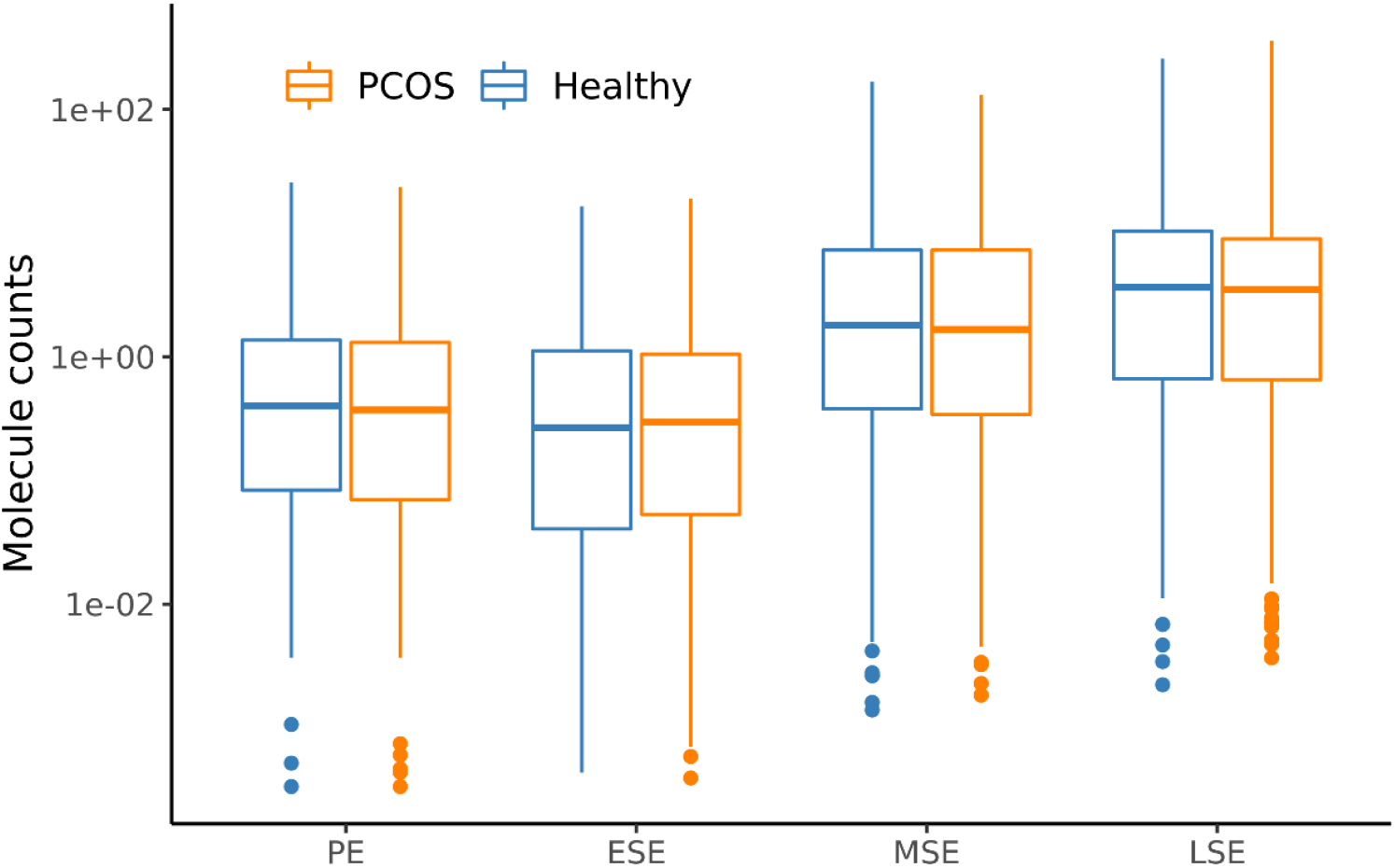
Gene expression profiles of endometrial biomarker genes in samples of PCOS patients and healthy women. The average gene expression (per group) of all assayed and housekeeper-normalised genes is presented for healthy and PCOS samples in the MD group. The lower and upper hinges of the boxplot correspond to the 25th and 75th percentiles. PE – proliferative phase, ESE – early-secretory phase, MSE – mid-secretory phase, LSE – late-secretory phase.

### beREADY model development

Before test development, 11 samples from the MD group were excluded from downstream analysis due to inconsistency between histology results and LH-day measurements. The remaining 63 MD group samples clustered clearly according to the menstrual cycle phases and enabled determining the receptivity class prediction (**Fig. 3**). When the PE phase and ESE samples were analysed together, the signature of receptivity genes allowed the discrimination of three groups of pre-receptive (PE and ESE samples), receptive (MSE samples), and post-receptive (LSE) samples (**Suppl. Fig. 3**). According to predicted menstrual cycle phase probabilities, the MSE endometria (n=17) were defined as receptive, corresponding to WOI time. PE and ESE endometrial samples (n=36) were classified as pre-receptive, suggesting that the endometrium has not yet reached the WOI. The LSE endometria (n=10) were defined as post-receptive, reflecting the period after WOI (**Fig. 4**). Based on 5-fold cross-validation, the predictive model classified PE/ESE, MSE, and LSE samples with an accuracy of 98.8%, averaged over all the receptivity classes.

**Figure 3.**
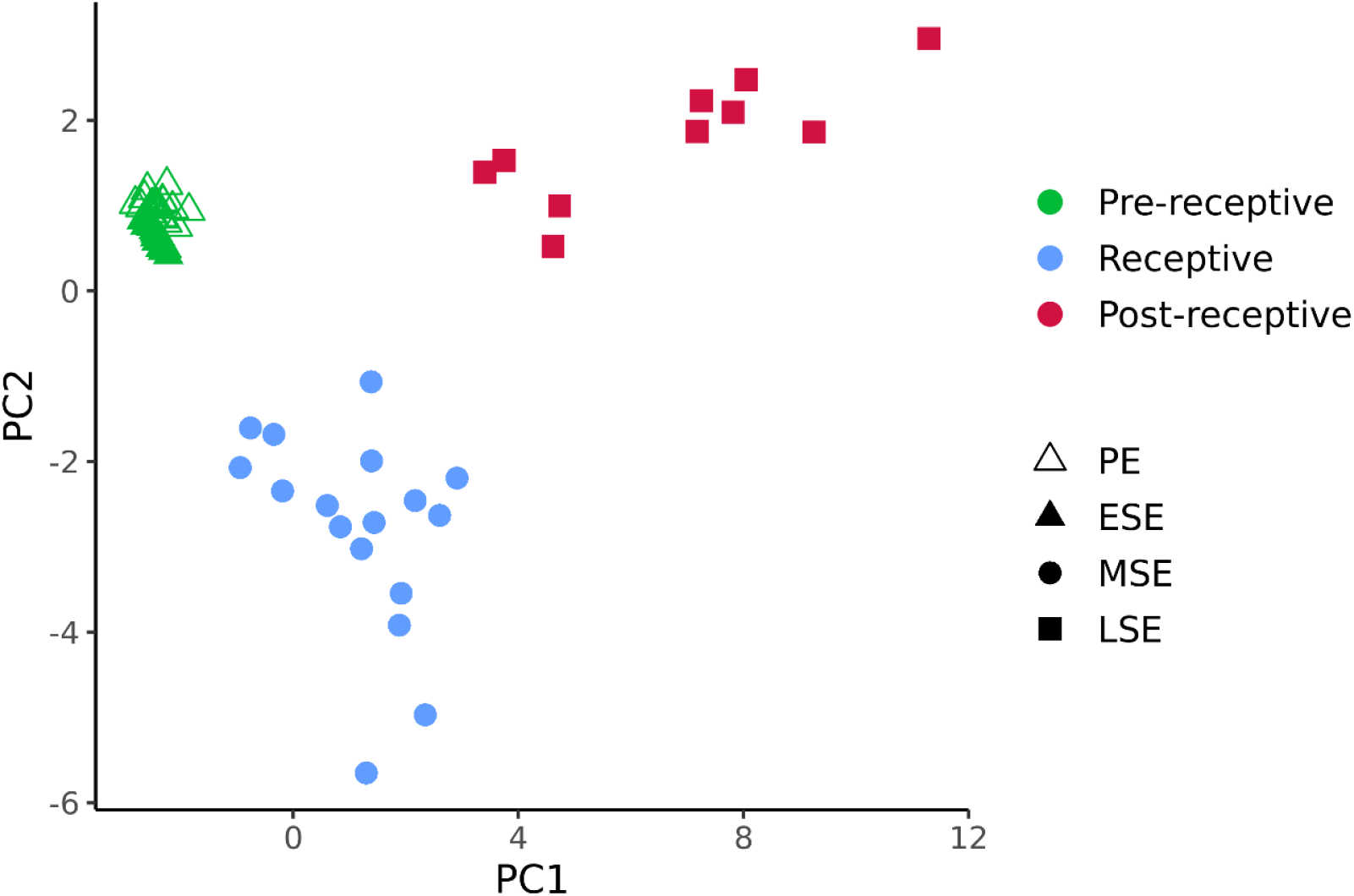
PCA plot of the model development group detects distinct transcriptional profiles of endometrial phases. The UMI-corrected counts were normalised with the geometric mean of housekeepers and scaled. PE – proliferative phase, ESE – early-secretory phase, MSE – mid-secretory phase, LSE – late-secretory phase.

**Figure 4.**
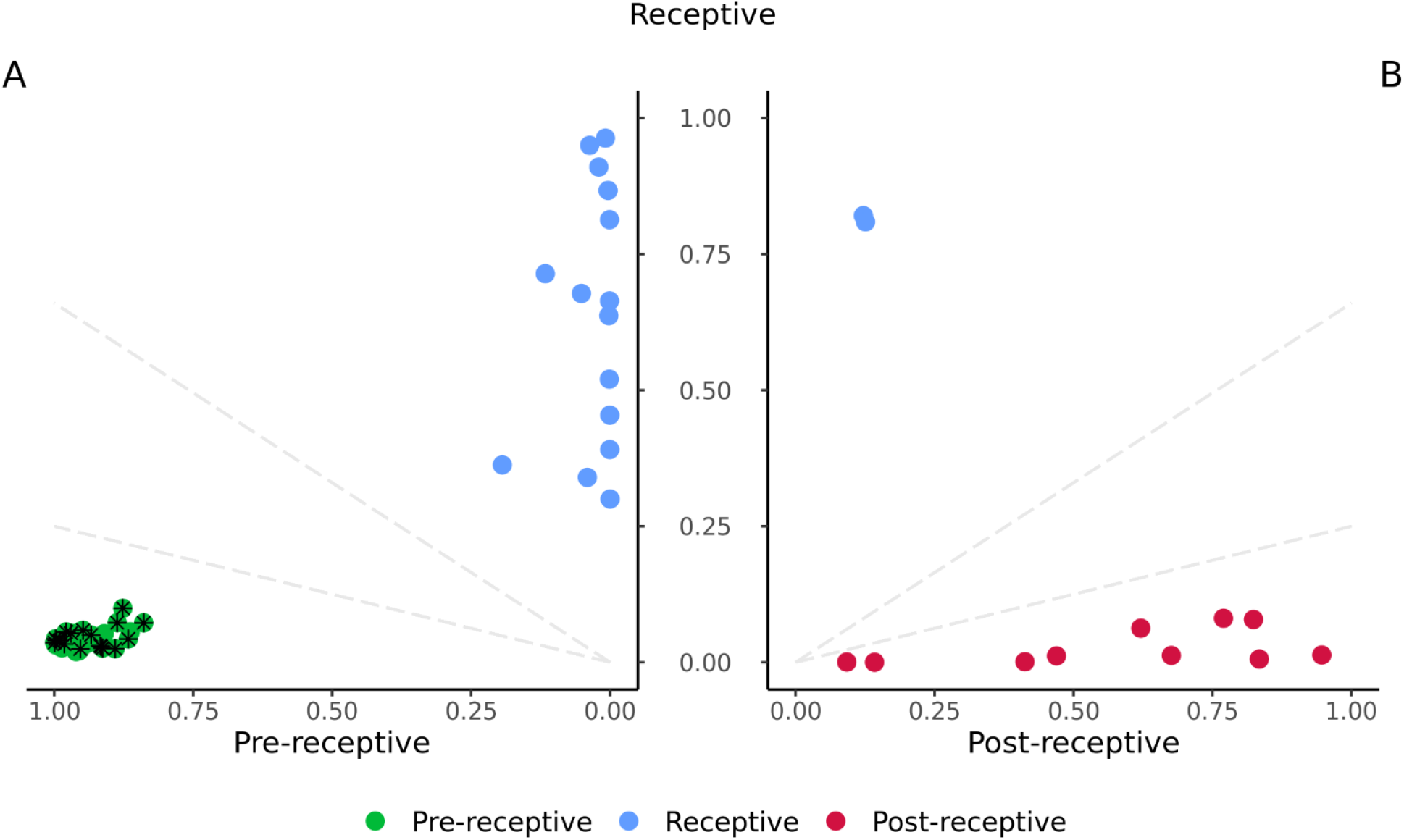
beREADY model output probabilities for model training and development group (MD) samples that belong to pre-receptive, receptive and post-receptive groups. The dashed lines represent intermediary groups as early-receptive and late-receptive decision boundaries, which are not detected in this analysis step. **(A)** The clustering between pre-receptive and receptive groups. Proliferative samples are marked with an asterisk. **(B)** The clustering between receptive and post-receptive groups.

Endometrial samples with two similar receptivity classes were classified as belonging to a transitionary class. Samples between the pre-receptive and receptive groups were defined as early-receptive and samples between receptive and post-receptive as late-receptive. However, samples positioned in the early-receptive, receptive, or late-receptive groups were collectively considered to represent the normal variability of WOI.

### beREADY test validation

Endometrial tissue samples obtained from healthy volunteers (MV group) were used for classification accuracy validation. In total, tissue samples from NC ESE (n=26), MSE (n=26), and LSE (n=5) phase were analysed (**Table 1**). The test evaluated all ESE samples as pre-receptive. Considering the MSE group, 25 samples were classified as receptive (25/26, 96.2%), from which six samples were classified as early-receptive (6/26, 23.1%) (**Table 2**). One sample from the MSE group (1/26, 3.8%) was classified as pre-receptive, concordant with prior histological evaluation. All LSE validation group samples were classified as post-receptive samples (**Figure 5**). In conclusion, all samples with concordant histological and LH dating were classified to the expected receptivity group, while one biopsy (1/57, 1.8%) demonstrated a discrepancy with the beREADY classification. It is relevant to note that a slight WOI shift, within the normal variability of the WOI range, was detected in almost every fourth healthy woman (23.1%) in MSE (**Table 2**).

**Table 2.**
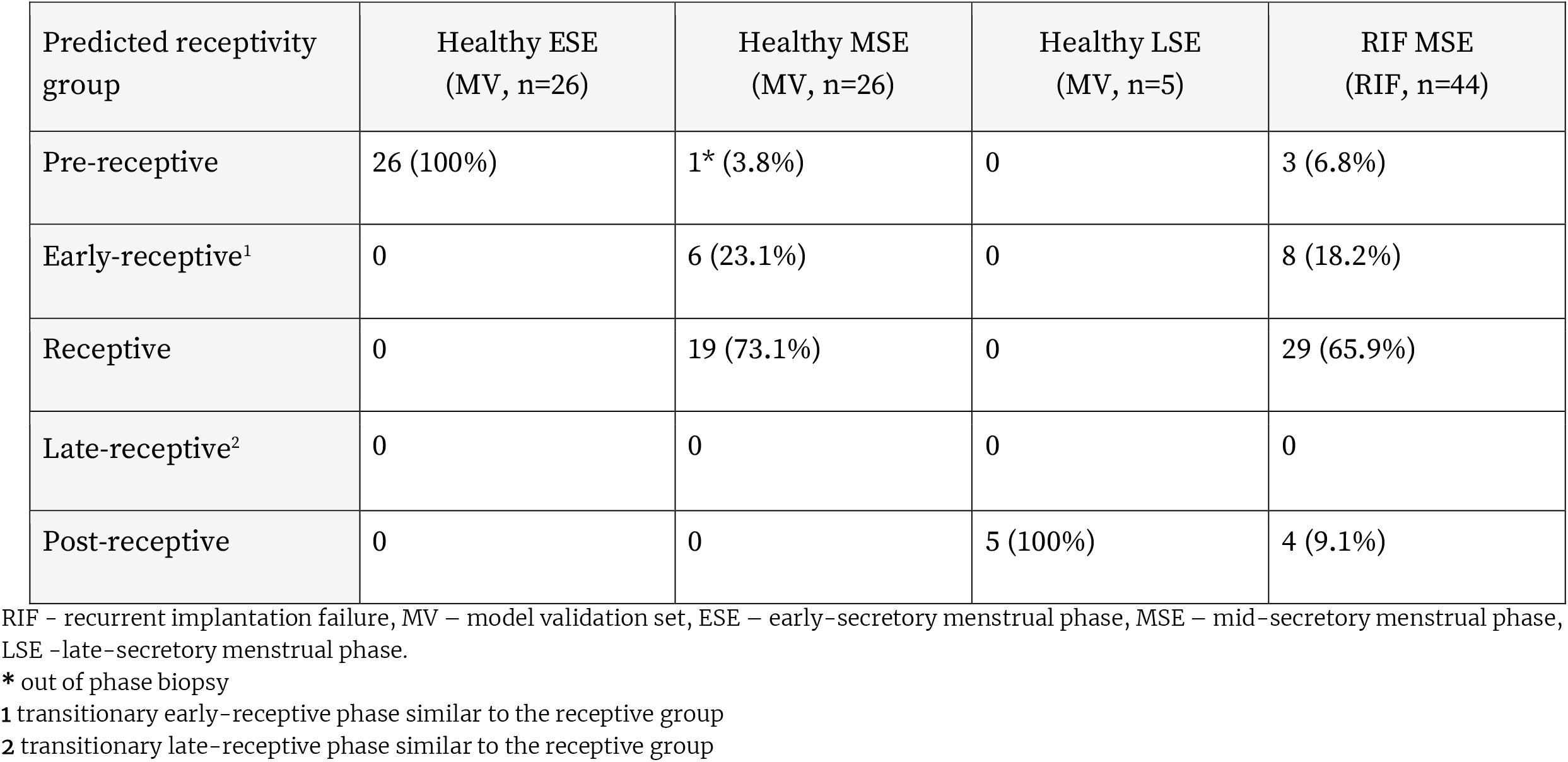
Endometrial receptivity status according to the beREADY test

**Figure 5.**
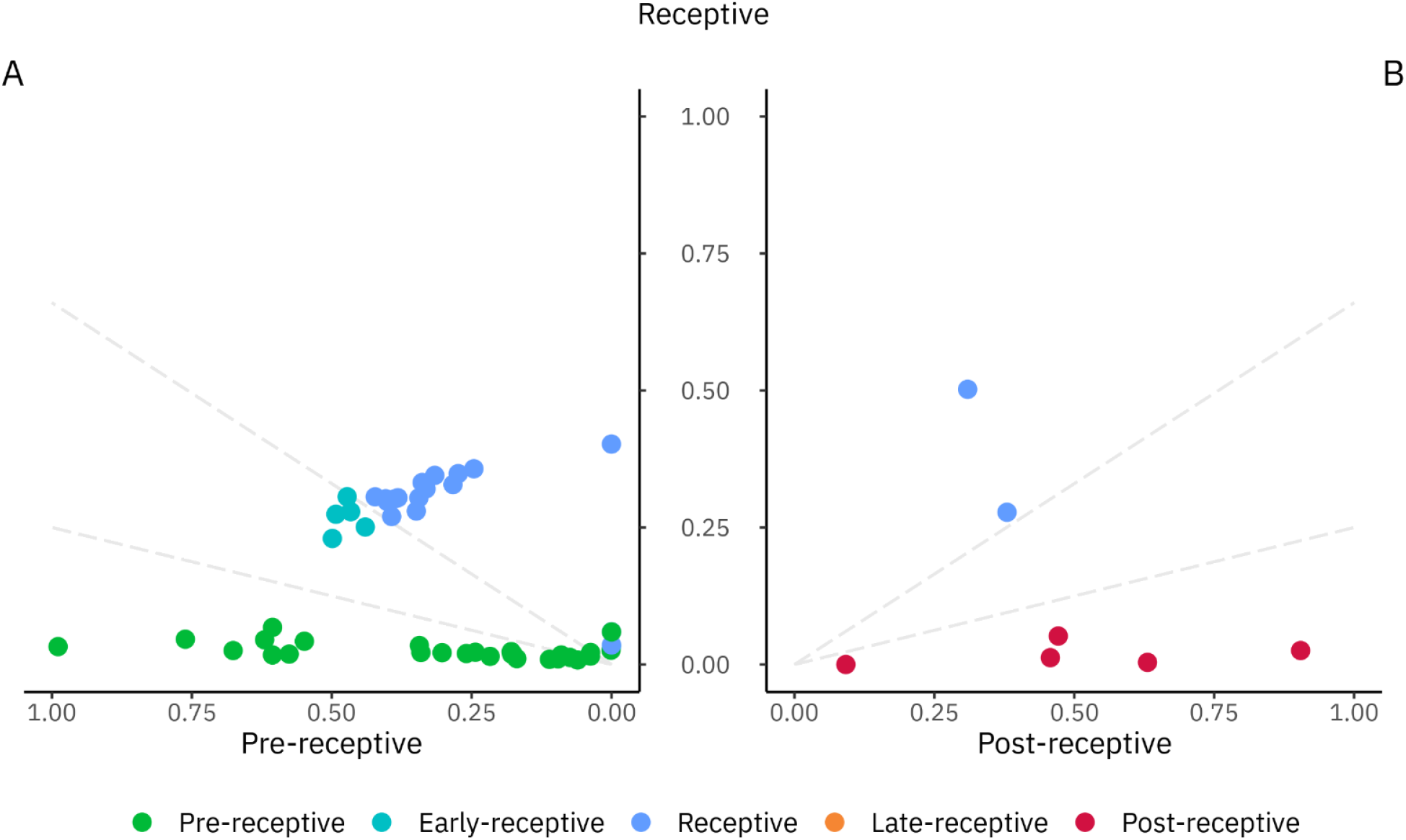
A focused analysis of the model validation (MV) samples, revealing early-receptive samples in mid-secretory group. Each point on this plot represents the probability of belonging to the given class. The colours represent the final prediction class. **(A)** Comparative pre-receptive and receptive group analysis revealed the intermediary early-receptive class shown in boundaries and positioned between dashed lines. **(B)** Pair-wise receptive and post-receptive group analysis confirmed five post-receptive samples shown in classification boundaries represented by dashed lines.

### RIF group study

Finally, a group of infertile women with RIF diagnosis (n=44) were analysed. Out of the RIF MSE samples, three patients (3/44, 6.8%) had a pre-receptive profile and displaced WOI (**Table 2**). No late-receptive samples were detected, but four samples displayed a post-receptive profile (4/44, 9.1%). Additionally, eight patients (8/44, 18.2%) were in the normal variability of the WOI range but deviated slightly from the receptive expressional profile and were classified as early-receptive. As a result, displaced WOI was detected in 15.9% of the samples in the RIF group. The proportion of the endometrial samples with the displaced WOI was higher in the RIF group than in the MV group, composed of healthy women, 15.9% and 1.8%, respectively (p=0.012).

To conclude, we present a novel TAC-seq based beREADY endometrial receptivity test, which enables targeted and highly sensitive PCR-bias free detection of endometrial receptivity transcriptomic biomarkers. As exemplified by an independent set of validation samples, the beREADY test allows accurate classification of endometrial receptivity of the studied samples, relevant for personalised WOI detection.

## Discussion

Previous transcriptomic studies of endometrium have shed light on the complex cross-talk mechanism between implanting embryos and the maternal uterus. This insight has explained why embryo implantation in some women repeatedly fails, regardless of the use of seemingly high-quality embryos [3, 14, 27]. Therefore, the focus on helping the RIF patients has shifted towards elucidating the maternal factors contributing to the unsuccessful IVF cycles. This interest has propagated the development of transcriptomic analysis tools for determining the receptivity status of the endometrium [11, 18, 20, 21]. We demonstrate that it is possible to find the personalised optimal WOI timing for embryo transfer with advanced sequencing technologies.

In this study, we present a novel endometrial receptivity detection method – the beREADY test. The test is based on a highly quantitative TAC-seq assay that allows precise and cost-effective endometrial receptivity biomarker analysis. A custom computational model for classifying sequenced samples was developed to analyse data generated by the TAC-seq pipeline. As it is based on the expressional profile of a targeted set of genes, the beREADY classification model can be possibly used in high-coverage whole-transcriptome studies. As a result, using endometrial biomarkers discovered from datasets with high predictive power for receptivity [23, 28], our prediction model allows the construction of the continuous transcriptomic timeline of endometrial receptivity development for personalised IVF and embryo transfer.

Several published and clinically available endometrial receptivity tests have been developed [11, 18, 20, 21]. Those tests use varying sets of targeted genes and different methodologies. The ERA test analyses 238 receptivity genes using massively parallel sequencing. Map®/ER Grade® test and ERPeakSM test analyse 40 genes, and WIN-Test 11 genes, respectively, by quantitative PCR. The selection of genes in the beREADY test is based on the comprehensive meta-analysis of endometrial receptivity biomarkers [23], complemented by eleven additional genes and four housekeeper genes. TAC-seq technology eliminates laboratory-caused PCR duplicates and counts the biomarkers at a single-molecule level [22]. To our knowledge, the beREADY test is the only endometrial receptivity test that applies UMI technology, enabling original transcript counts estimation while avoiding the PCR-caused bias in results. This approach has already been used to determine the menstrual cycle phases from endometrial tissue [29].

For beREADY test development, we used natural cycle endometrial samples from healthy women and women with PCOS diagnoses. Although the involvement of endometrial factors in PCOS-associated infertility has been suggested (reviewed [13]), there are no large-scale and systematic studies simultaneously analysing numerous endometrial receptivity-related genes in different cycle phases in PCOS patients. In addition, endometrial tissues of PCOS patients have demonstrated altered response to steroids [30, 31] but the overall effect of PCOS on pregnancy outcome is still unclear [32]. Therefore, we tested if PCOS alters the transcriptomic profiles of our selection of receptivity biomarkers. We found no significant differences between healthy controls and PCOS patients in the expression profile of endometrial biomarkers throughout the menstrual cycle from PE to LSE phase. Based on these findings, we concluded that the selected receptivity biomarkers are unaffected in women with PCOS, and the samples were suitable for use as a reference together with samples from healthy women in the development of beREADY model. As a limitation, our study analysed PCOS patients who were neither obese nor had oligomenorrhea. When combined, PCOS and obesity can still introduce an additional risk factor for impaired endometrial receptivity [33, 34]. For these reasons, our conclusions regarding receptivity in PCOS patients are valid for women with PCOS patients within the normal range of BMI.

We validated the beREADY test by analysing endometrial biopsies from healthy women representing the fertile female population, displaying a high similarity between sample collection time and the molecular test result (56/57, 98.2%). This similarity can be explained by the fact that the healthy women in our validation group were all LH tested and had ultrasound confirmation of ovulation and typical hormonal values. Despite this, one MSE sample (1/26, 3.8%) with morphology suggesting the PE phase had shifted WOI according to the beREADY test. In that case, the beREADY test classified the sample as pre-receptive, suggesting that the WOI had not yet arrived. However, displaced WOI among oocyte donors and women undergoing IVF without RIF diagnosis has been previously reported [9, 35]. In the MSE group, we also classified six samples as early-receptive (6/26, 23.1%), suggesting a slight shift in WOI that remained within the normal range of receptivity. Therefore, transcriptomic profiling can offer additional accuracy for determining the individual receptivity timing of the endometrium for embryo transfer. This information can potentially be considered when fine-tuning the timing of the personalised embryo transfer.

The criteria for RIF diagnosis are controversial and include several factors, such as the number of failed treatment cycles, number of transferred good quality embryos and maternal age [36]. Due to its heterogeneous nature, the causes of RIF have remained largely unknown. When focusing solely on endometrial factors, it has been proposed by Sebastian-Leon et al. that RIF is caused by at least two distinct molecular phenomena of displaced (asynchronous) or disrupted (pathological) WOI [8]. In our study, we applied the beREADY test to detect the rate of displaced WOI in a study group of 44 RIF patients. In total, we detected shifted WOI in 15.9% of RIF cases, which is slightly less than the results described previously by Lessey et al., 25% [6], Mahajan et al., 27.5% [37], Hashimoto et al., 24% [10], and Patel et al. 17.7% [27] in women with RIF diagnosis. Compared with the MV group, the RIF study group demonstrated a significantly higher proportion of shifted WOI cases (p<0.05), indicating that displaced WOI can cause RIF in those cases. Nevertheless, the rate of displaced WOI in RIF is present merely in a sixth of the cases, suggesting additional factors, such as embryo-related and those disrupting the WOI, play an essential role in the aetiology of RIF.

Although the MD set consisted of the samples with clearly different expressional profiles corresponding to the different menstrual cycle phases, this study suffers from a relatively small sample size with uneven distribution between the receptivity classes. Furthermore, to estimate the clinical usefulness of the model, an extensive randomised controlled trial (RCT) must be carried out among the RIF patients, comparing the implantations, pregnancy, and live birth rates between pET and conventional embryo transfer cycles. Ideally, the RCT should be carried out with euploid embryos following preimplantation genetic testing for aneuploidy (PGT-A), which could rule out the genetic defects of IVF embryos as one of the possible causes of implantation failure. PGT-A has been shown to improve the cumulative IVF pregnancy outcome, avoiding the RIF diagnosis in some patients because of the transfer of the euploid embryos only [38]. Therefore, in our study, the RIF samples with possibly embryo-associated implantation failure were not excluded, likely explaining our findings that receptive endometrium was found in ca 80% of RIF cases.

Embryonic and maternal factors such as embryonal chromosomal aberrations, impaired endometrial receptivity, maternal immune dysfunction, and infertility-associated diseases, like endometriosis and male factors, can all affect the chance for embryo implantation. A large-scale retrospective study involving nearly 4,500 patients undergoing IVF with PGT-A discovered that only around 5% of the infertile patients failed to achieve the clinical pregnancy following three consecutive frozen euploid single embryo transfers. This outcome suggests that the rate of actual recurrent implantation failure may be lower [38]. Nevertheless, other studies have demonstrated a significant increase in implantation rate with pET without applying PGT-A. In a study by Simon et al., receptivity testing before the first embryo transfer increases the implantation and pregnancy rate of first embryo transfer and the cumulative pregnancy rate when the pET is compared with the conventional embryo transfers [19]. Another study by Haouzi et al. demonstrated that the implantation rate increases over three times, from 7% to 23%, after using the personalised WOI determination in IVF patients with RIF diagnosis [3]. Therefore, the potential utility of personalised endometrial dating in IVF may increase the chance of implantation of the first IVF treatment and improve the treatment outcome of the patients with two or more previous failed IVF cycles.

In conclusion, our results demonstrate that TAC-seq can be successfully applied for transcriptomic endometrial dating. This assay detected distinct profiles of endometrial samples at pre-receptive, receptive, and post-receptive stages. Additionally, we found no significant difference in the expression levels of receptivity biomarkers in the healthy fertile women and women diagnosed with PCOS. Based on these findings, we developed the beREADY endometrial receptivity test with a custom classification model for distinguishing between the transcriptional profiles. In our tests, beREADY detected an increased rate of displaced WOI cases in the RIF study group compared to the validation group. Consequently, these findings suggest that applying the beREADY test before embryo transfer could potentially reduce the chance of implantation failure rate in the RIF patients.

## Data Availability

All data produced in the present study are available upon reasonable request to the authors.

## Acknowledgements

This research was funded by the Estonian Research Council (grant PRG1076), Horizon 2020 innovation grant (ERIN, grant no. EU952516), the EU-FP7 Marie Curie Industry-Academia Partnerships and Pathways (IAPP, grant SARM, EU324509), the Enterprise Estonia (grant no. EU48695) and by Polish Ministry of Health (grant no. 6/6/4/1/NPZ/2017/1210/1352).

## Supplementary Tables

**Supplementary Table 1.**
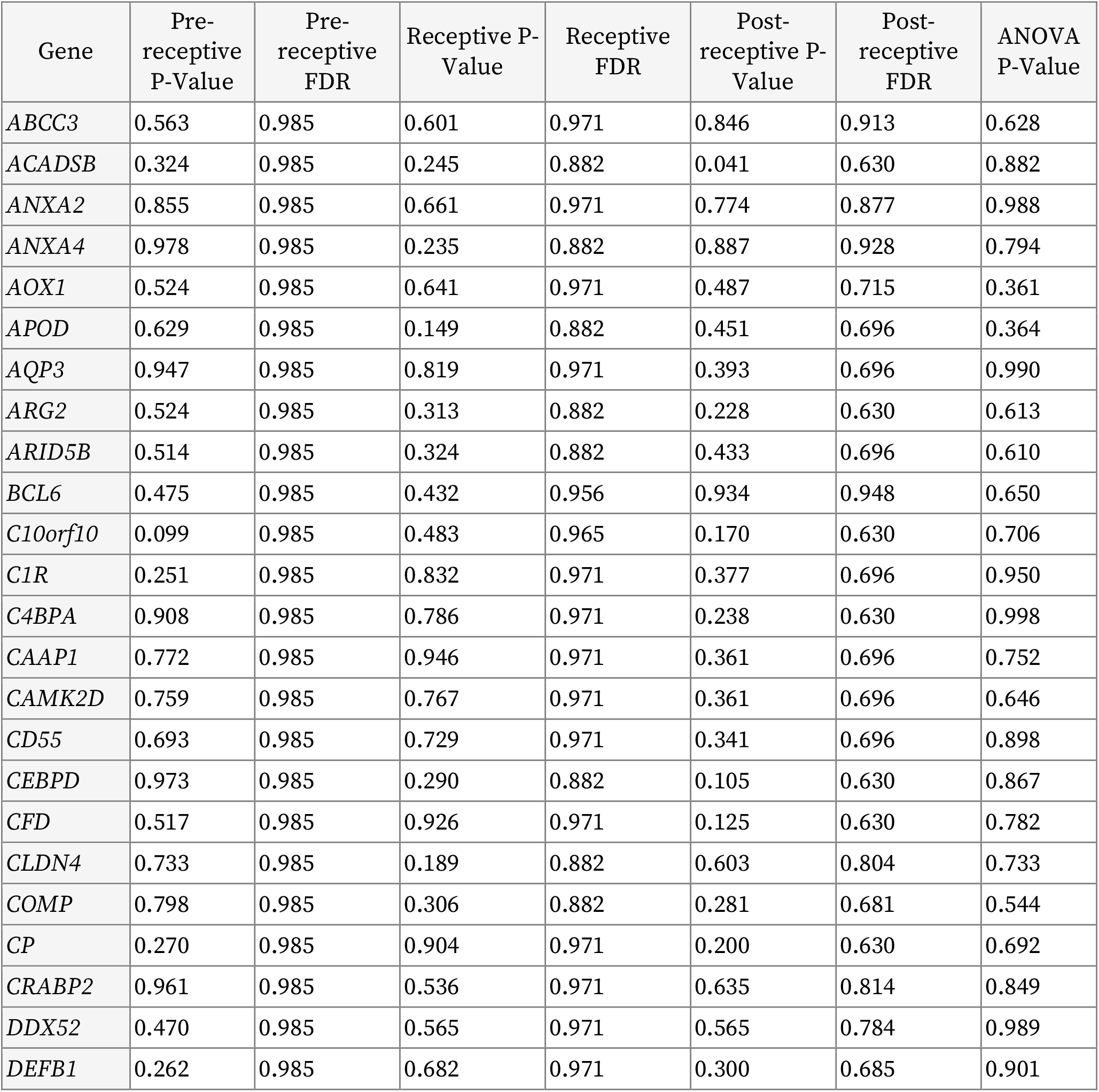
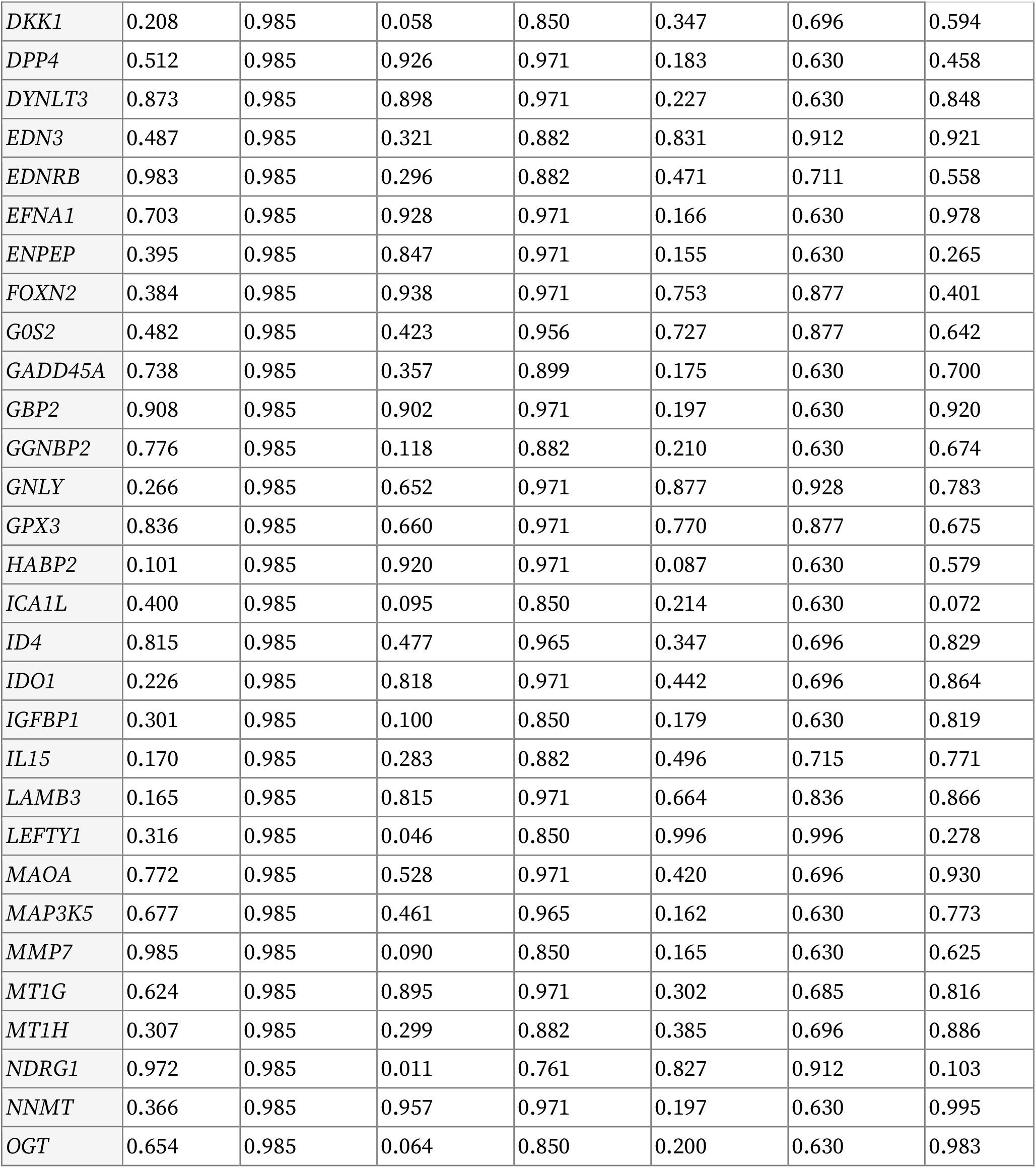
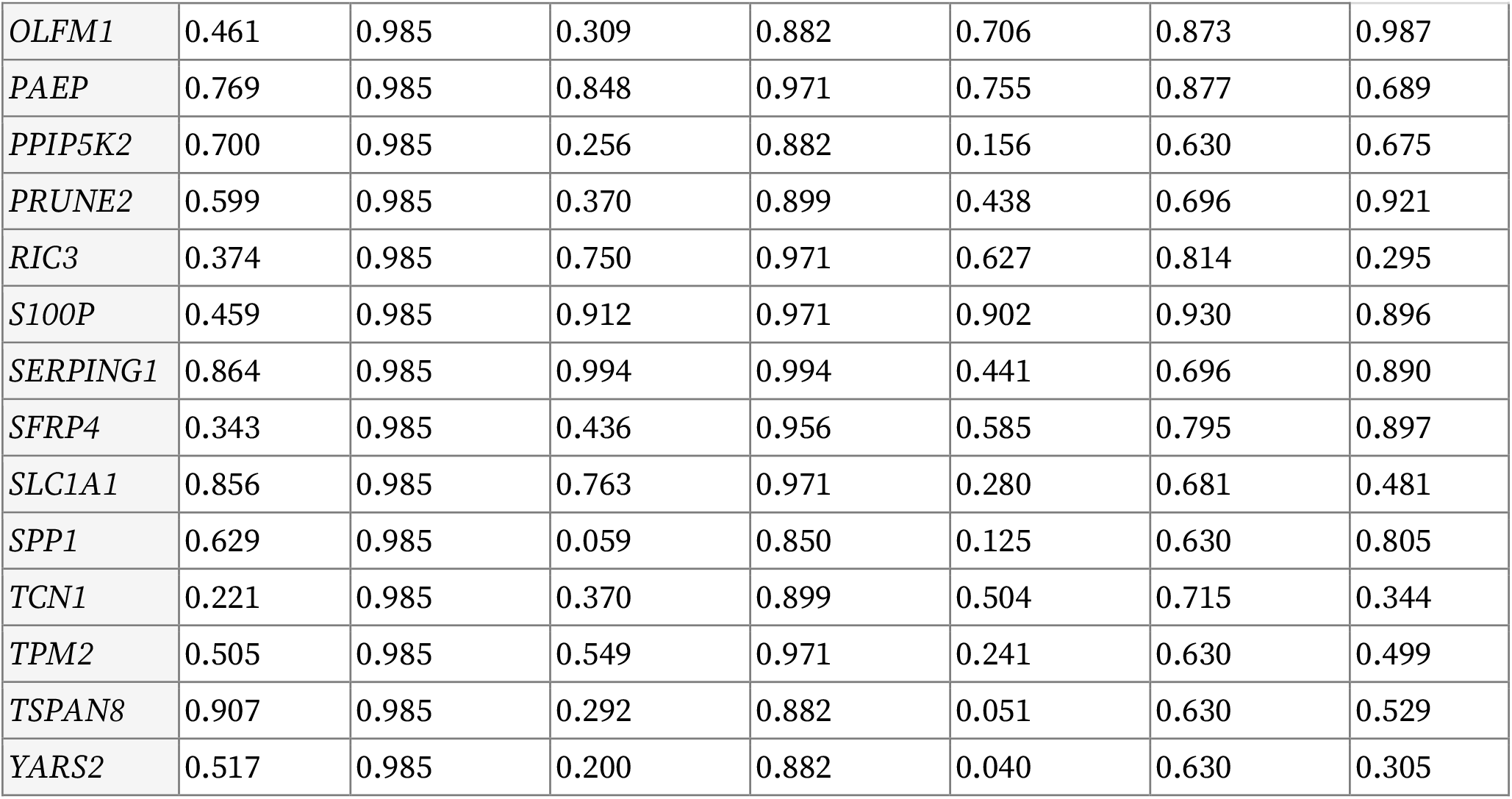
Gene-wise differential analysis between PCOS and healthy subjects. FDR – false discovery rate; ANOVA – analysis of variance.

## Supplementary Figures

**Supplementary Figure 1.**
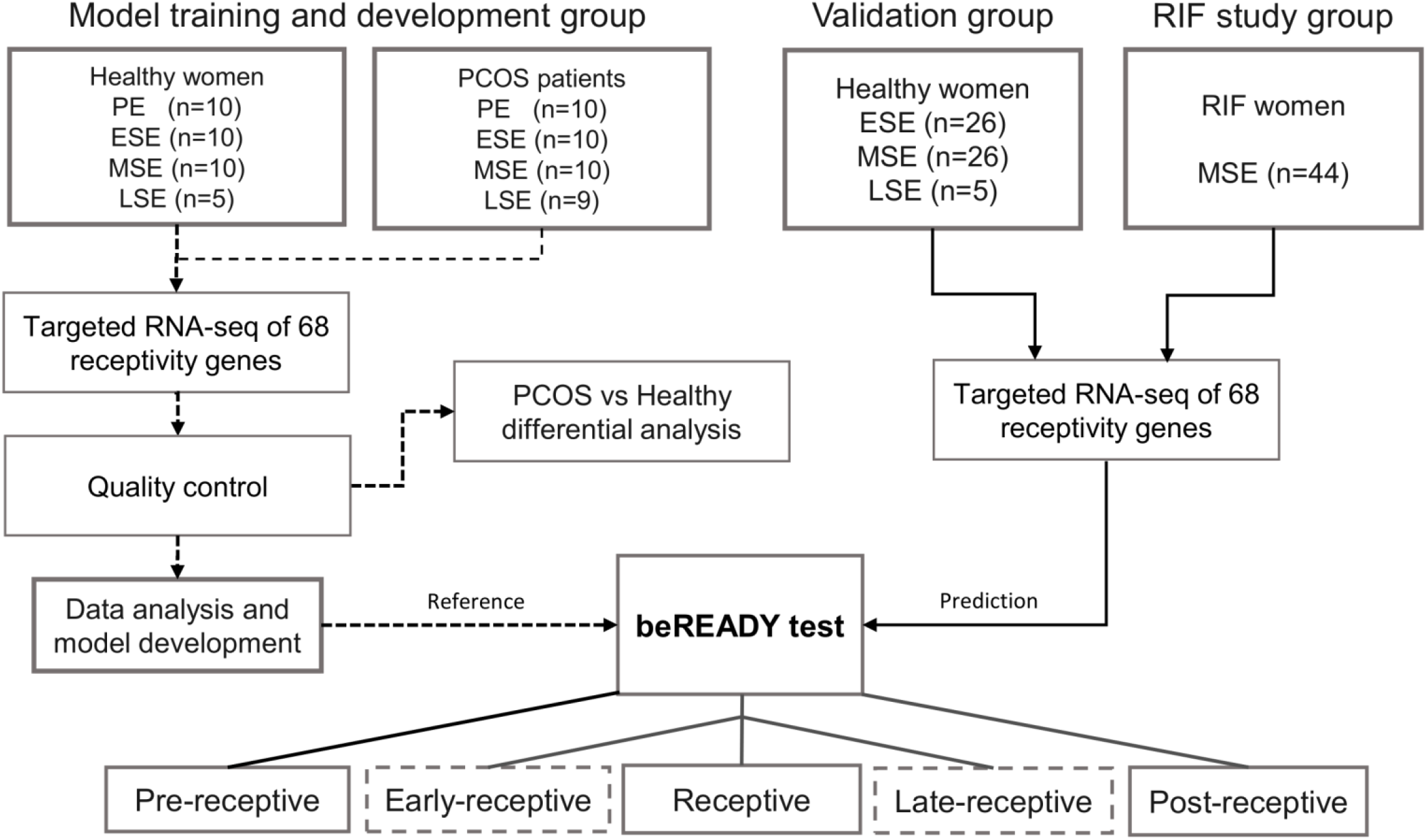
Overview of the study design. RIF – recurrent implantation failure, PCOS – polycystic ovary syndrome, ESE – early-secretory phase, MSE – mid-secretory phase, LSE – late-secretory phase.

**Supplementary Figure 2.**
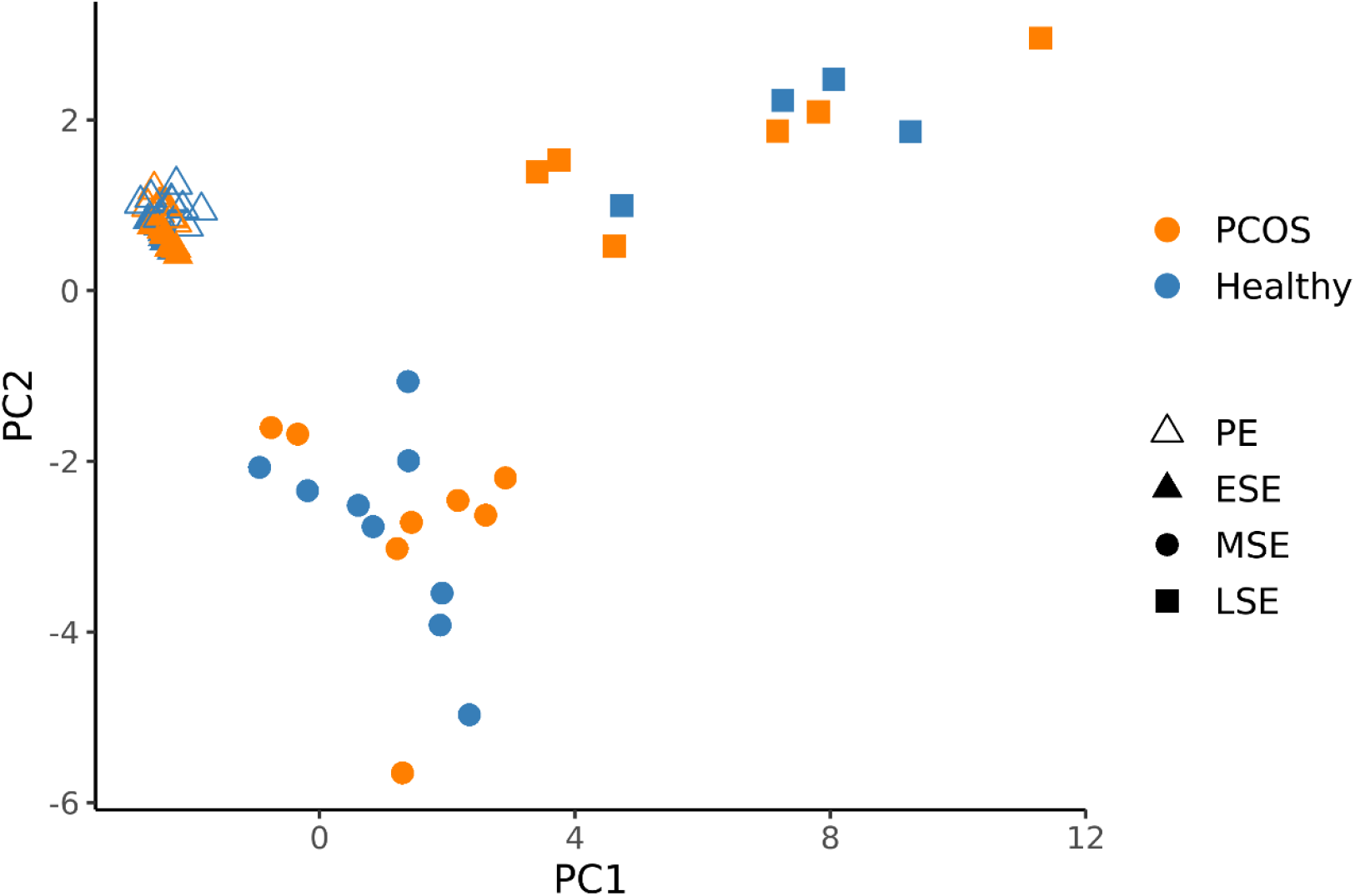
PCA plot of the model development (MD) set. The UMI-corrected counts were normalised with the geometric mean of housekeepers and scaled. PCOS – polycystic ovarian syndrome, PE – proliferative phase, ESE – early-secretory phase, MSE – mid-secretory phase, LSE – late-secretory phase.

**Supplementary Figure 3.**
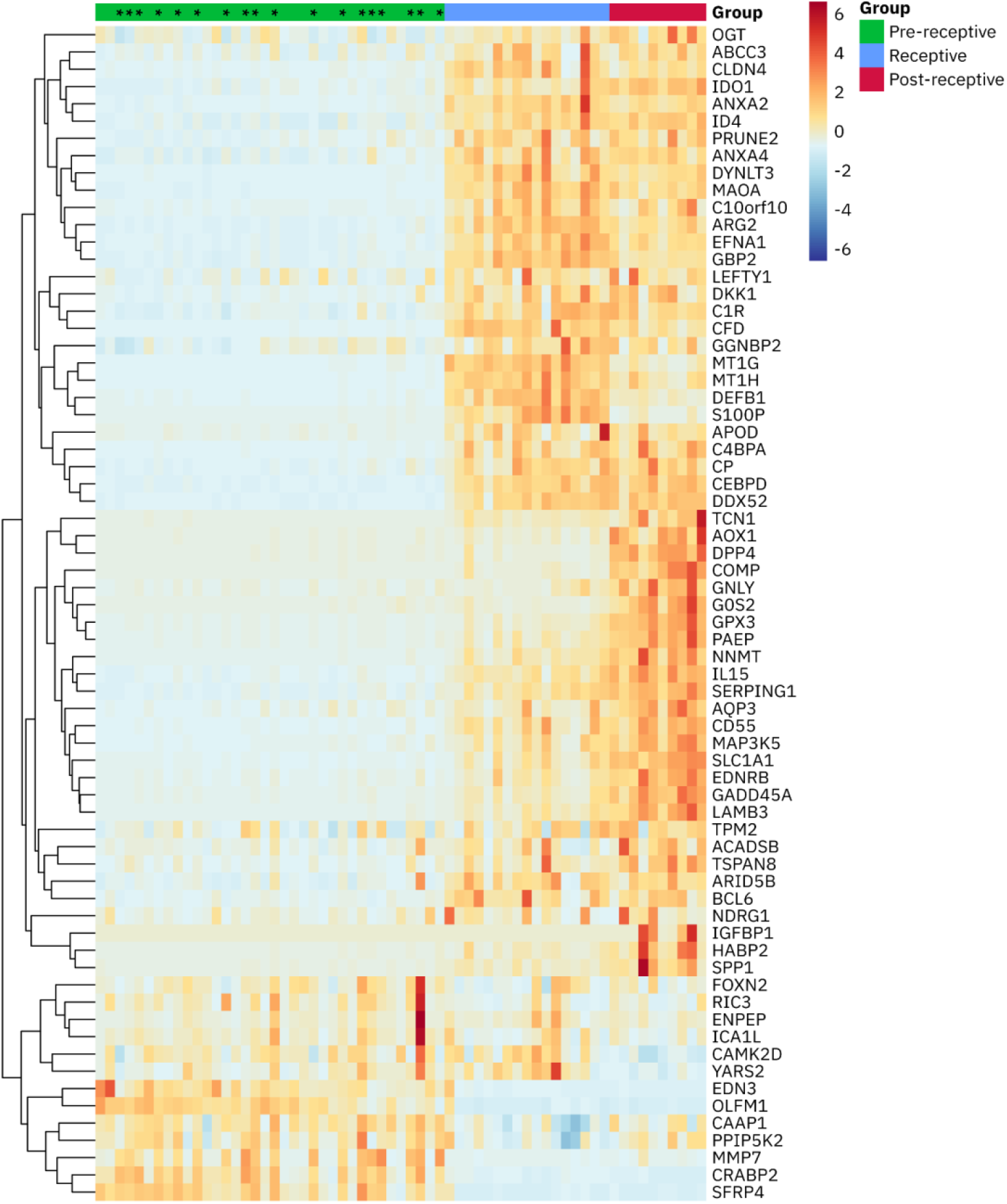
Heatmap of the scaled expression levels of normalised model training and development group samples. Samples are ordered by the output score of the model. The genes are clustered hierarchically. Proliferative samples are marked with an asterisk (*).

